# Mental health and health behaviours before and during the initial phase of the COVID-19 lockdown: Longitudinal analyses of the UK Household Longitudinal Study

**DOI:** 10.1101/2020.06.21.20136820

**Authors:** Claire L Niedzwiedz, Michael J Green, Michaela Benzeval, Desmond D Campbell, Peter Craig, Evangelia Demou, Alastair H Leyland, Anna Pearce, Rachel M Thomson, Elise Whitley, S Vittal Katikireddi

**Affiliations:** Institute of Health & Wellbeing, University of Glasgow; MRC/CSO Social & Public Health Sciences Unit, University of Glasgow; Institute for Social and Economic Research, University of Essex; Public Health Scotland

## Abstract

**Background:** There are concerns that COVID-19 mitigation measures, including the “lockdown”, may have unintended health consequences. We examined trends in mental health and health behaviours in the UK before and during the initial phase of the COVID-19 lockdown and differences across population subgroups.

**Methods:** Repeated cross-sectional and longitudinal analysis of the UK Household Longitudinal Study, including representative samples of adults (aged 18+) interviewed in four survey waves between 2015 and 2020 (n=48,426). 9,748 adults had complete data for longitudinal analyses. Outcomes included psychological distress (General Health Questionnaire-12 (GHQ)), loneliness, current cigarette smoking, use of e-cigarettes and alcohol consumption. Cross-sectional prevalence estimates were calculated and multilevel Poisson regression assessed associations between time period and the outcomes of interest, as well as differential associations by age, gender, education level and ethnicity.

**Results:** Psychological distress increased one month into lockdown with the prevalence rising from 19.4% (95% CI 18.7%-20.0%) in 2017-19 to 30.3% (95% CI 29.1%-31.6%) in April 2020 (RR=1.3, 95% CI: 1.1,1.4). Groups most adversely affected included women, young adults, people from an Asian background and those who were degree educated. Loneliness remained stable overall (RR=0.9, 95% CI: 0.6,1.5). Smoking declined (RR=0.9, 95% CI=0.8,1.0) and the proportion of people drinking four or more times per week increased (RR=1.4, 95% CI: 1.3,1.5), as did binge drinking (RR=1.5, 95% CI: 1.3,1.7).

**Conclusions:** Psychological distress increased one month into lockdown, particularly among women and young adults. Smoking declined, but adverse alcohol use generally increased. Effective measures are required to mitigate adverse impacts on health.

What is already known on this topic
- Countries around the world have implemented radical COVID-19 lockdown measures, with concerns that these may have unintended consequences for a broad range of health outcomes.
- Evidence on the impact of lockdown measures on mental health and health-related behaviours remains limited.

What this study adds
- In the UK, psychological distress markedly increased during lockdown, with women particularly adversely affected.
- Cigarette smoking fell, but adverse drinking behaviour generally increased.

## Introduction

The coronavirus disease 2019 (COVID-19) pandemic has led to large-scale societal changes in many countries. Governments have introduced substantial restrictions to people’s movement, including limiting potential to attend work and school, or see friends and family.^1 2^ Such ‘lockdown’ measures could have large impacts on health and health inequalities.^3 4^ While some impacts could arise from reduced access to healthcare during lockdown^5^, lockdown measures themselves could have direct consequences on mental health and health-related behaviours.

Research prior to the pandemic has suggested quarantine is linked to several negative psychological outcomes.^6^ During the COVID-19 pandemic, concerns have been repeatedly raised about potentially long-lasting harms to mental health.^7^ Similarly, health-related behaviours such as alcohol consumption and smoking could be subject to rapid change in either direction. Increased stress during lockdown could increase consumption,^8 9^ while greater awareness of health risks, reduced availability and socialising could reduce consumption.

The UK Government introduced strict physical distancing measures, or ‘lockdown’ on the 23^rd^ March 2020, with other mitigation measures being introduced throughout March (Appendix 1 Box S1).^10^ This restricted the general population to staying at home, unless required to leave for the purposes of carrying out an essential job (referred to as a ‘keyworker’, such as transport, education, food and health and social care workers), to buy necessary items or to take exercise.

Understanding the impact of lockdown is important as further periods of physical distancing are likely to be necessary in many countries for some time, especially as the possibility of further waves of infection remain. These impacts may disproportionately affect specific population subgroups, with concerns that young people, women and disadvantaged socioeconomic groups may be at greater risk. We investigated the impact of the UK’s COVID-19 lockdown on mental health and health behaviours, as well as whether any observed impacts differed by age, gender, ethnicity and education level.

## Methods

### Data source

The UK Household Longitudinal Study (also referred to as ‘Understanding Society’) is a nationally representative longitudinal household panel study, based on a clustered stratified probability sample of UK households, described in detail previously.^11^ All adults (aged 16+ years) in chosen households are invited to participate. Data collection for each ‘wave’ usually spans 24 months, with participants re-interviewed annually by online, face-to-face or telephone survey. We used pre-pandemic data from wave 7 (2015-2017), wave 8 (2016-18) and wave 9 (2017-19), with household response rates of 81-84%.^11 12^ Following the pandemic’s onset, an additional wave of data was collected via online survey between 24^th^ and 30^th^ April 2020 (referred to as the COVID-19 wave A – henceforth the ‘CA wave’).^13^ The response rate for the CA wave was 48.6% of those who took part at wave 9.^14 15^ We analysed data from all adults aged 18+ years who participated in each wave for repeated cross-sectional analysis (excluding proxy interviews). When analysing educational inequalities we restricted analyses to adults aged 25+ years as educational attainment tends to be stable from that age onwards.^16^ For longitudinal analysis, we included participants with complete data from all four waves and aged 18+ years during wave 9.

The University of Essex Ethics Committee has approved all data collection for the Understanding Society main study and COVID wave. No additional ethical approval was necessary for this secondary data analysis.

### Outcomes

Mental health was assessed at all four waves using the General Health Questionnaire-12 (GHQ-12), which is a screening tool for psychological distress that has been validated for use in epidemiological studies.^17^ Respondents scoring 4 or more (out of a possible total of 12) are likely to be experiencing anxiety and/or depression.^18 19^ To better understand the driving symptoms of any change in psychological distress we also considered each individual GHQ item in subsidiary analyses, investigating trends in the proportion of respondents who selected the two most adverse response categories for each question. We also conducted sensitivity analyses with the item on enjoyment of day-to-day activities removed (since this could be affected by lockdown restrictions without necessarily indicating poor mental health), and with the cut-off point reduced to 3 or more symptoms, as a way of examining increases in less severe psychological distress and to enable comparison with other studies using this definition. Loneliness was assessed at wave 9 and the CA wave by asking participants: “in the last 4 weeks, how often did you feel lonely?” and respondents were able to answer hardly ever or never, some of the time, or often. In the statistical models this was converted to a binary variable (often felt lonely versus all other responses).

We also assessed three health behaviour outcomes: cigarette smoking; e-cigarette use; and alcohol consumption. Participants were asked “do you smoke cigarettes? Please do not include electronic cigarettes (e-cigarettes)” and those who answered ‘yes’ were then asked, “approximately how many cigarettes a day do you usually smoke, including those you roll yourself?”. We defined cigarette smoking (excluding e-cigarettes) as current smoker versus non-smoker and the number of cigarettes per day was calculated (<10,10-19, 20+ cigarettes per day) for subsidiary analyses. Current e-cigarette use was defined on the basis of having used e-cigarettes at least once a week (waves 8, 9 and CA wave). Information about alcohol consumption was collected (waves 7, 9 and CA wave) using the Alcohol Use Disorders Identification Test for Consumption (AUDIT-C) instrument.^20^ However, the CA wave contained some modifications including asking about drinking behaviour over the last four weeks, rather than the last year. We therefore looked at three key outcomes: binge drinking (6+ drinks in a single sitting on weekly basis), frequency of alcohol consumption (4+ times per week) and heavy drinking (5+ drinks on a typical day when drinking).

### Covariates

We adjusted for a range of potential confounders that were likely causes of the outcomes and that did not lie on the causal pathway between lockdown and the outcomes: age group (18-24, 25-44, 45-64, 65+ years at wave 9) and self-reported gender (male/female). Highest education level, coded as: degree-level or equivalent qualifications, A-level/AS-level or equivalent, General Certificate of Secondary Education (GCSE) or equivalent, and no qualifications. Race/ethnicity was categorised as: white, Asian, black, mixed, and other, but recoded to binary (white and non-white) for the statistical models due to small numbers within specific ethnic minority groups. Interview year as a continuous variable accounted for temporal trends.

### Statistical analysis

Prevalence estimates (with 95% confidence intervals) for each outcome were calculated in repeated cross-sectional analyses using all complete sets of responses from waves 7 to 9 and the CA wave. Cross-sectional inverse probability weights provided with the data were used to adjust for attrition and to create estimates that were representative of the general population over time (see Appendix 1 for details of the weights). This was supplemented with additional weighting for differences in outcome non-response by age, gender, ethnicity and education. We repeated cross-sectional analyses stratified by gender, age group, ethnicity and education level.

We then restricted our sample to individuals with repeated measures for all relevant waves for longitudinal analysis (n=9,748). We conducted multi-level Poisson regression with robust standard errors, to assess associations between outcomes and the time period an observation was taken in (CA wave or prior), adjusting for age, gender, race/ethnicity, and interview year. Poisson regression was used to calculate relative risks.^21^ Robust standard errors were used to improve the accuracy of estimated 95% confidence intervals and p-values given the data are clustered. We carried out a complete case analysis, using longitudinal inverse probability weights constructed for these models to adjust for attrition and missing data (see Appendix 1 for further details). We tested for differential associations by fitting interaction terms for age group, gender, educational level and race/ethnicity. Statistical analysis was conducted using Stata/MP 15.1 and R version 3.6.0 for the figures.

## Results

Table 1 describes the 48,426 individuals included in the repeated cross-sectional analysis by wave, after excluding participants with missing data (see Figure S1 for STROBE diagram and Table S1 for details of the longitudinal sample). The sample at the CA wave in April 2020 was: 53.4% female, 40.5% were degree level educated, 8.3% were from ethnic minority groups and the mean age of participants was 49.6 (95% CI: 49.0-50.1). Weighted prevalence estimates for the key outcomes before and during the COVID-19 pandemic are shown in Figure 1 and Table S2 (Appendix 2 contains prevalences for each wave and outcome by subgroup).

**Table 1:**
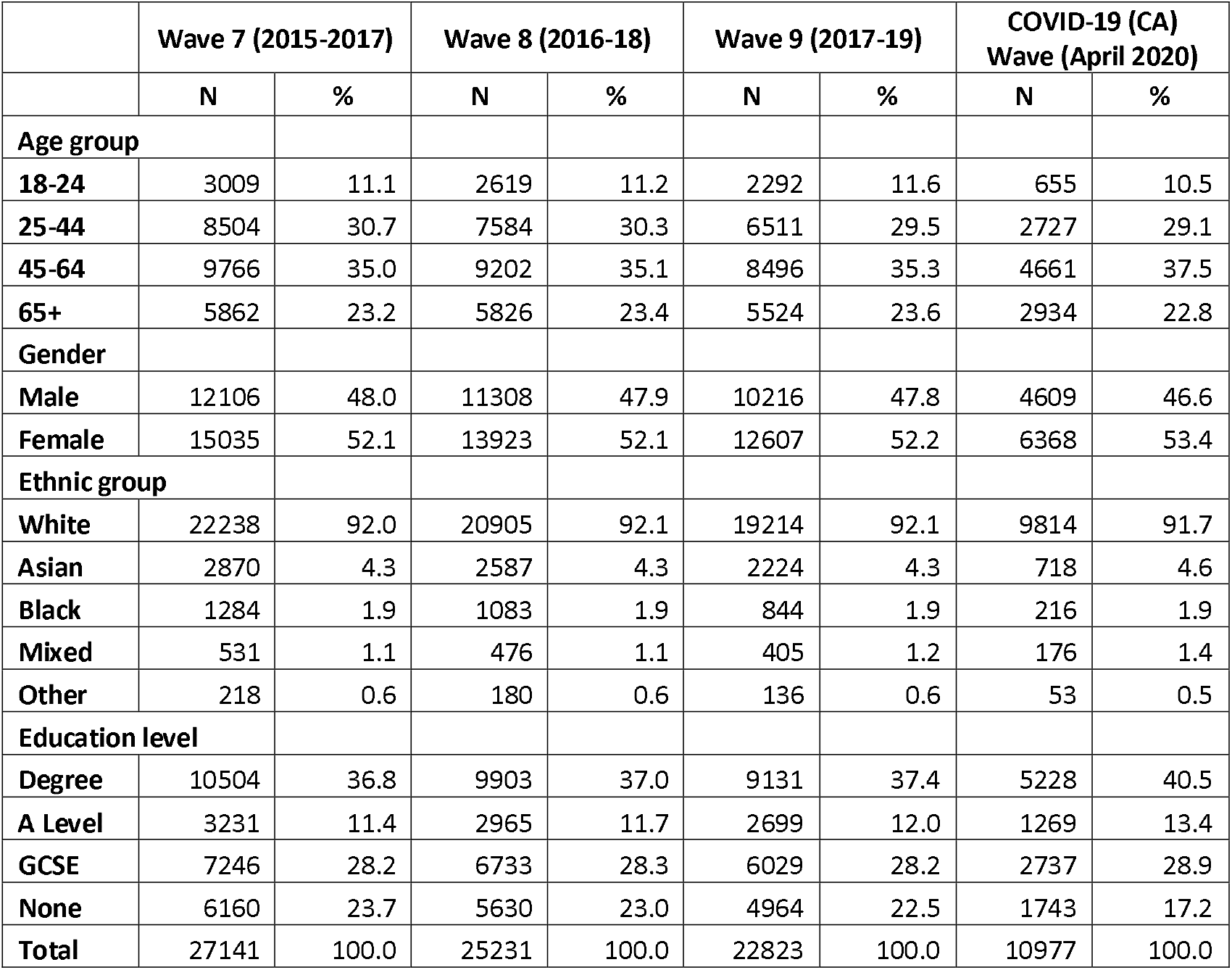
Description of the repeated cross-sectional samples (weighted %)

|  | Wave 7 (2015-2017) |  | Wave 8 (2016-18) |  | Wave 9 (2017-19) |  | COVID-19 (CA)<br>Wave (April 2020) |  |
| --- | --- | --- | --- | --- | --- | --- | --- | --- |
|  | N | % | N | % | N | % | N | % |
| **Age group** |  |  |  |  |  |  |  |  |
| **18-24** | 3009 | 11.1 | 2619 | 11.2 | 2292 | 11.6 | 655 | 10.5 |
| **25-44** | 8504 | 30.7 | 7584 | 30.3 | 6511 | 29.5 | 2727 | 29.1 |
| **45-64** | 9766 | 35.0 | 9202 | 35.1 | 8496 | 35.3 | 4661 | 37.5 |
| **65+** | 5862 | 23.2 | 5826 | 23.4 | 5524 | 23.6 | 2934 | 22.8 |
| **Gender** |  |  |  |  |  |  |  |  |
| **Male** | 12106 | 48.0 | 11308 | 47.9 | 10216 | 47.8 | 4609 | 46.6 |
| **Female** | 15035 | 52.1 | 13923 | 52.1 | 12607 | 52.2 | 6368 | 53.4 |
| **Ethnic group** |  |  |  |  |  |  |  |  |
| **White** | 22238 | 92.0 | 20905 | 92.1 | 19214 | 92.1 | 9814 | 91.7 |
| **Asian** | 2870 | 4.3 | 2587 | 4.3 | 2224 | 4.3 | 718 | 4.6 |
| **Black** | 1284 | 1.9 | 1083 | 1.9 | 844 | 1.9 | 216 | 1.9 |
| **Mixed** | 531 | 1.1 | 476 | 1.1 | 405 | 1.2 | 176 | 1.4 |
| **Other** | 218 | 0.6 | 180 | 0.6 | 136 | 0.6 | 53 | 0.5 |
| **Education level** |  |  |  |  |  |  |  |  |
| **Degree** | 10504 | 36.8 | 9903 | 37.0 | 9131 | 37.4 | 5228 | 40.5 |
| **A Level** | 3231 | 11.4 | 2965 | 11.7 | 2699 | 12.0 | 1269 | 13.4 |
| **GCSE** | 7246 | 28.2 | 6733 | 28.3 | 6029 | 28.2 | 2737 | 28.9 |
| **None** | 6160 | 23.7 | 5630 | 23.0 | 4964 | 22.5 | 1743 | 17.2 |
| **Total** | 27141 | 100.0 | 25231 | 100.0 | 22823 | 100.0 | 10977 | 100.0 |

**Figure 1:**
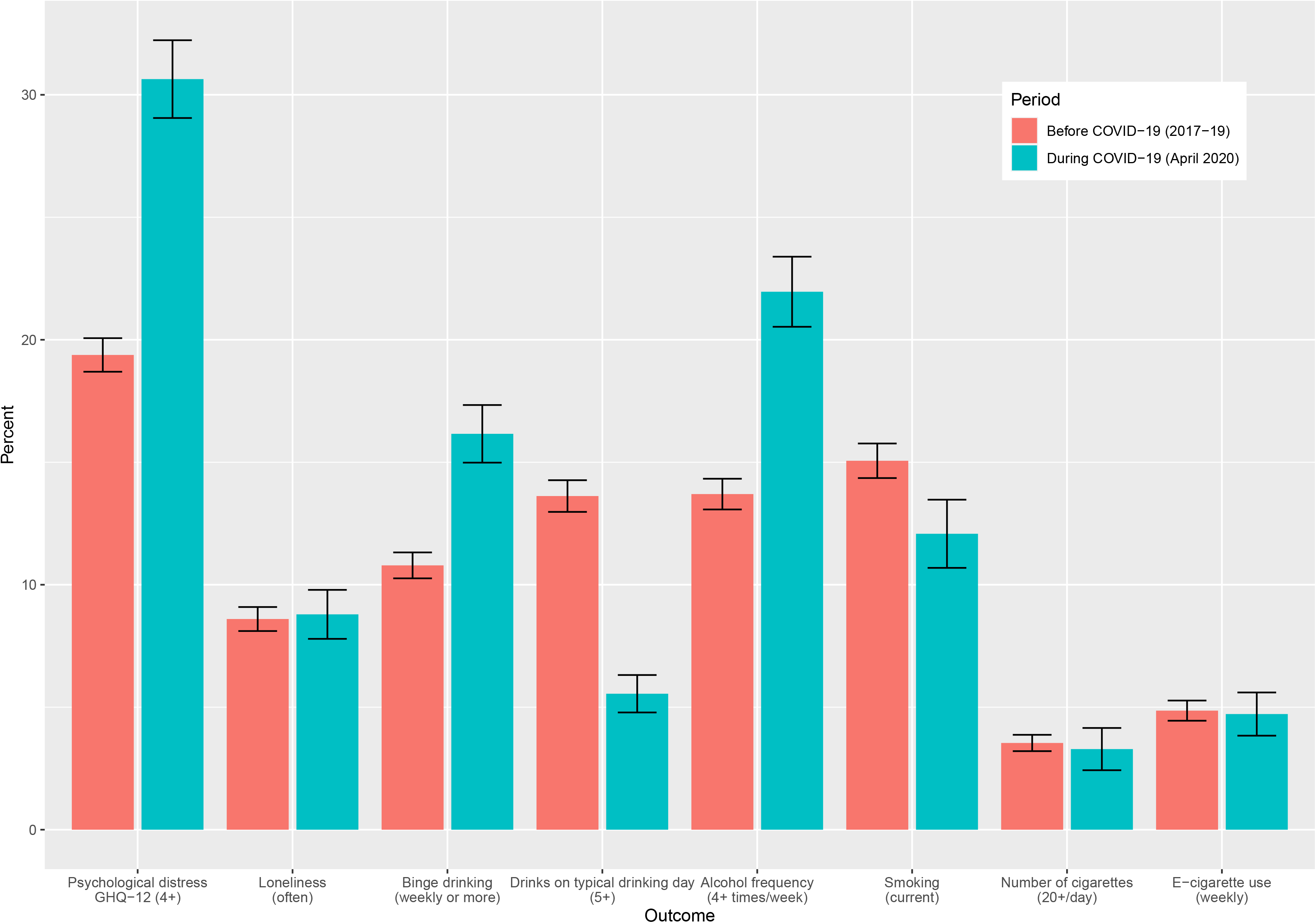
Mental health and health behaviours before (2017-2019) and during the COVID-19 lockdown (April 2020)

### Psychological distress

Psychological distress has steadily increased over time from 17.6% (95% CI: 17.0-18.2) in 2015-17 (wave 7) to 19.4% (95% CI: 18.7-20.1) in 2017-19 (wave 9), but substantially increased to 30.3% (95% CI: 29.1-31.6) during the COVID-19 pandemic in April 2020 (Figure 1 and Table S2). All symptoms of psychological distress worsened over this period (Figure 2). The symptom which had the largest deterioration was enjoyment of normal day-to-day activities. Worsening symptoms were also observed for concentration, sleep, feelings of unhappiness and loss of purpose. In contrast, there was less of an apparent increase in feelings of worthlessness, an inability to overcome difficulties and lacking confidence. In sensitivity analyses using 3+ symptoms as the cut-off point, the prevalence of psychological distress increased from 23.7% (95% CI: 23.0-24.5) in 2017-19 (wave 9) to 37.8% (95% CI: 36.5-39.2) in the CA wave (Table S2). We also investigated whether the decline in enjoyment of day-to-day activities was driving the increase in psychological distress. Removing this item reduced the magnitude of the increase, but it remained substantial (Table S2).

**Figure 2:**
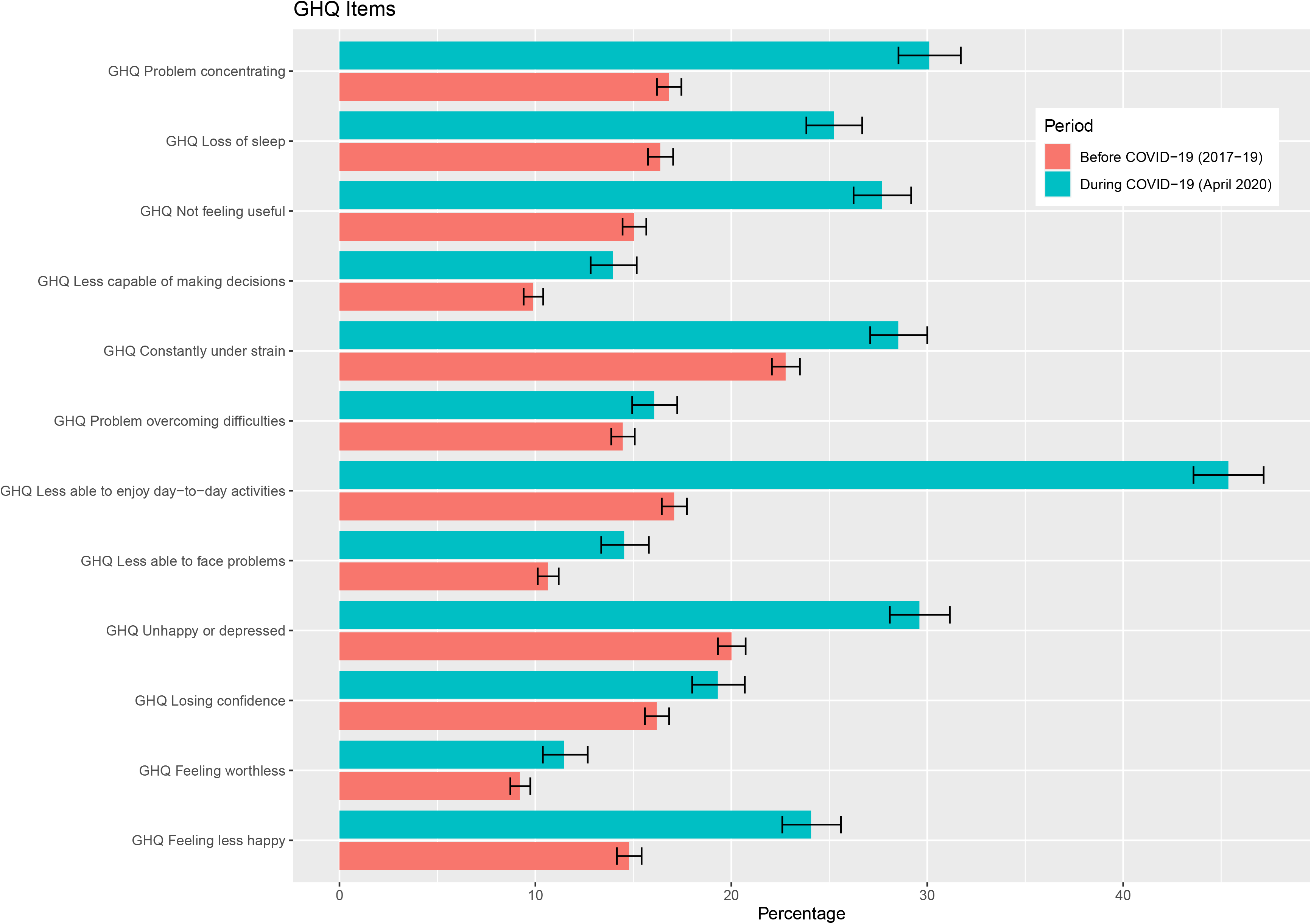
Psychological distress (General Health Questionnaire items) before (2017-2019) and during the COVID-19 lockdown (Apri I 2020)

The increase in psychological distress was most pronounced among people aged under 45 years, as well as among the most educated groups (Figure 3 and Appendix 2). Women were also more adversely affected than men; among women the prevalence of psychological distress increased from 23.0% (95% CI: 22.0-23.9) in 2017-19 to 36.7% (95% CI: 35.1-38.4) during the pandemic period. Asian minority ethnic groups also experienced a large increase in psychological distress; from 18.7% (95% CI: 16.4-21.2) to 36.1% (95% CI: 30.7-41.9) (Appendix 2).

**Figure 3:**
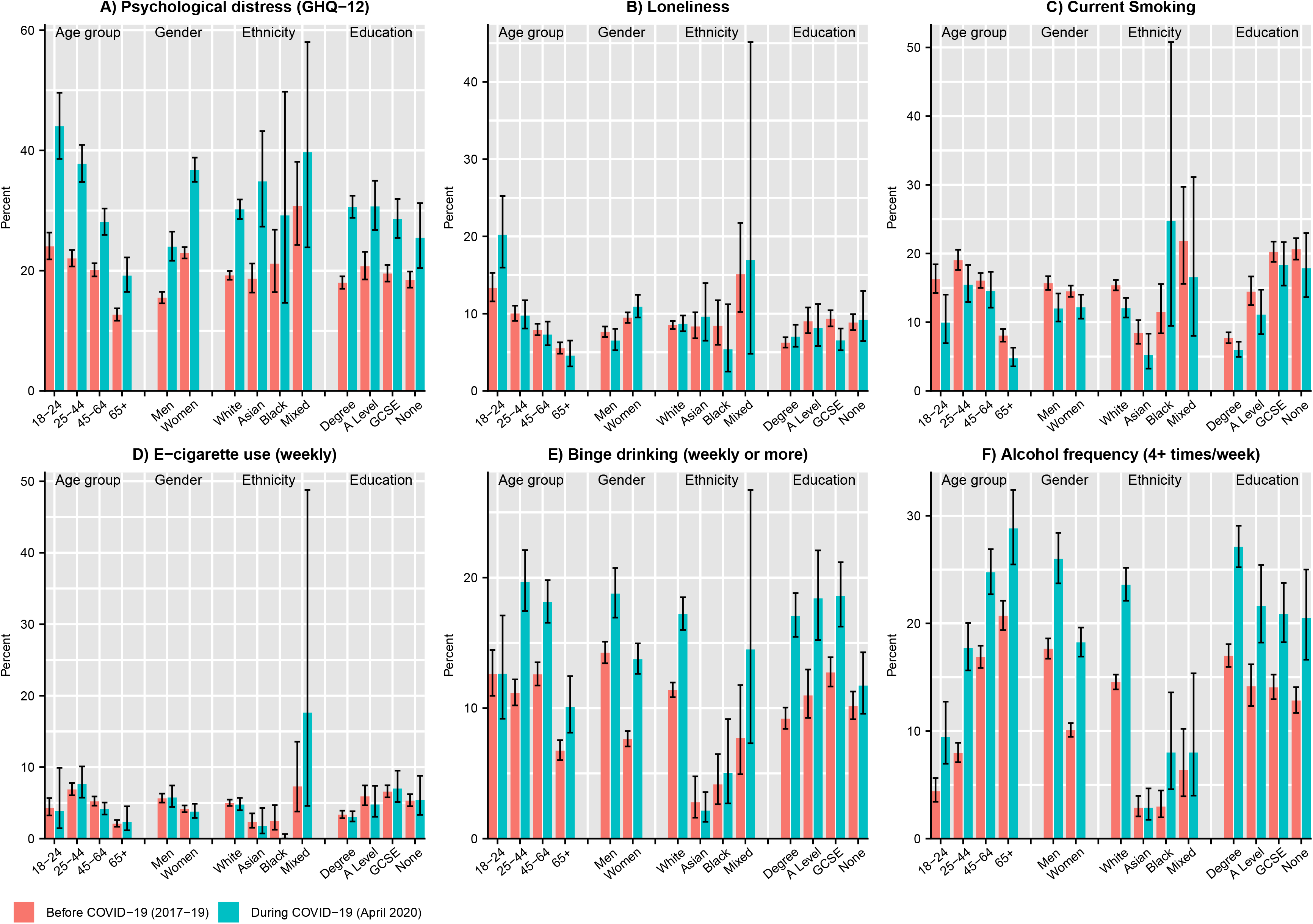
Mental health and health behaviours before (2017-2019) and during the COVID-19 lockdown (April 2020) by subgroup

Longitudinal regression models (Table 2) adjusted for age, gender, race/ethnicity and interview year demonstrated that the risk of psychological distress was elevated during the pandemic compared with the pre-pandemic period (RR: 1.3, 95% CI: 1.2-1.4), taking into account prior trends. In sensitivity analyses using the lower cut-off threshold for GHQ the RR was 1.4 (95% CI: 1.3-1.5). There was evidence of differential effects by age group, gender and education level when examining statistical interactions with time period (Table S3).

**Table 2:** Risk ratios (RR) derived from the multilevel Poisson models for mental health and health behaviour outcomes.

|  | Psychological distress (GHQ-12, 4+ cut-off) | Psychological distress (GHQ-12, 3+ cut-off) | Loneliness | Binge drinking (weekly or more) | Alcohol frequency (4+ times per week) | 5+ drinks on typical drinking day | Current cigarette smoking | Regular e-cigarettes |
| --- | --- | --- | --- | --- | --- | --- | --- | --- |
|  | RR [95% CI] | RR [95% CI] | RR [95% CI] | RR [95% CI] | RR [95% CI] | RR [95% CI] | RR [95% CI] | RR [95% CI] |
| **Interview year** | 1.06<sup>***</sup><br>[1.03,1.09] | 1.05<sup>***</sup><br>[1.02,1.07] | 1.00<br>[0.83,1.20] | 1.02<br>[0.98,1.06] | 1.05<sup>***</sup><br>[1.03,1.08] | 1.02<br>[0.98,1.06] | 0.99<br>[0.97,1.01] | 1.11<sup>*</sup><br>[1.00,1.22] |
| **Age group: 18-24** | 2.33<sup>***</sup><br>[2.03,2.67] | 2.08<sup>***</sup><br>[1.85,2.34] | 4.76<sup>***</sup><br>[3.55,6.39] | 1.59<sup>***</sup><br>[1.25,2.01] | 0.17<sup>***</sup><br>[0.13,0.23] | 15.29<sup>***</sup><br>[11.73,19.93] | 2.85<sup>***</sup><br>[1.62,5.02] | 2.04<br>[0.95,4.39] |
| 25-44 | 2.01<sup>***</sup><br>[1.83,2.22] | 1.82<sup>***</sup><br>[1.68,1.98] | 2.73<sup>***</sup><br>[2.15,3.46] | 1.73<sup>***</sup><br>[1.50,2.01] | 0.48<sup>***</sup><br>[0.43,0.54] | 7.62<sup>***</sup><br>[6.06,9.59] | 6.56<sup>***</sup><br>[4.42,9.75] | 6.23<sup>***</sup><br>[4.14,9.37] |
| 45-64 | 1.71<sup>***</sup><br>[1.55,1.88] | 1.54<sup>***</sup><br>[1.43,1.67] | 2.02<sup>***</sup><br>[1.61,2.54] | 1.98<sup>***</sup><br>[1.74,2.26] | 0.87<sup>**</sup><br>[0.79,0.95] | 4.46<sup>***</sup><br>[3.55,5.62] | 5.29<sup>***</sup><br>[3.56,7.87] | 3.94<sup>***</sup><br>[2.70,5.74] |
| 65+ (ref) |  |  |  |  |  |  |  |  |
| **Gender: Men (ref)** |  |  |  |  |  |  |  |  |
| Women | 1.58<sup>***</sup><br>[1.48,1.69] | 1.51<sup>***</sup><br>[1.43,1.60] | 1.64<sup>***</sup><br>[1.41,1.91] | 0.53<sup>***</sup><br>[0.49,0.59] | 0.62<sup>***</sup><br>[0.57,0.67] | 0.52<sup>***</sup><br>[0.46,0.59] | 1.00<br>[0.82,1.22] | 0.76<sup>*</sup><br>[0.58,0.99] |
| **Race/ethnicity: White (ref)** |  |  |  |  |  |  |  |  |
| Non-white | 1.14<sup>*</sup><br>[1.02,1.27] | 1.08<br>[0.98,1.19] | 1.12<br>[0.88,1.44] | 0.27<sup>***</sup><br>[0.20,0.36] | 0.34<sup>***</sup><br>[0.26,0.43] | 0.30<sup>***</sup><br>[0.22,0.40] | 1.19<br>[0.88,1.61] | 0.52<sup>*</sup><br>[0.32,0.86] |
| **Period: Pre-COVID-19** |  |  |  |  |  |  |  |  |
| During COVID-19 | 1.28<sup>***</sup><br>[1.15,1.42] | 1.38<sup>***</sup><br>[1.25,1.51] | 0.90<br>[0.55,1.47] | 1.48<sup>***</sup><br>[1.27,1.73] | 1.38<sup>***</sup><br>[1.26,1.51] | 0.40<sup>***</sup><br>[0.33,0.47] | 0.89<sup>**</sup><br>[0.82,0.97] | 0.66<sup>*</sup><br>[0.48,0.91] |
| **Observations** | 38992 | 38992 | 19496 | 29244 | 29244 | 29244 | 38992 | 29244 |

95% confidence intervals in brackets
\* $p < 0.05$ , \*\* $p < 0.01$ , \*\*\* $p < 0.001$

### Loneliness

Overall, loneliness remained relatively stable before and during the lockdown period (Figure 1). However, in repeated cross-sectional analysis, there were differences by age group (Figure 3), with younger people experiencing higher overall levels of loneliness, as well as a large increase in loneliness (from 13.3% (95% CI: 11.6-15.3) to 21.0% (95% CI: 17.2-25.5)) during lockdown. Loneliness also slightly increased among women, but fell among men. In longitudinal analyses, differences by age were less apparent (although this analysis had less statistical power), but there was evidence for an interaction between gender and time period (Table S4).

### Alcohol consumption

Binge drinking increased from 10.8% (95% CI: 10.3-11.3) in wave 9 (2017-19) to 16.5% (95% CI: 15.6-17.6) during lockdown (Figure 1), as did the proportion of people reporting drinking four or more times a week (13.7% (95% CI: 13.1-14.3) to 22.9% (95% CI: 21.7-24.1)). Differences by age group and gender were apparent. Binge drinking remained stable in the youngest age group but increased in those aged 25 and over (Figure 3). Binge drinking and frequent drinking also increased more among women, white people and the degree educated group.

The proportion of people reporting drinking five or more drinks during a typical day when drinking decreased from 13.6% (95% CI: 13.0-14.3) during wave 9 (2017-19) to 5.6% (95% CI: 4.8-6.4) during the pandemic lockdown (Figure 1). This decrease was marked in the youngest age group, falling from 31.9% (95% CI: 29.5-34.5) during wave 9 to 8.5% (95% CI: 5.4-13.2) during lockdown (Figure 3).

Results from longitudinal models supported cross-sectional analyses (Table 2), with the risk of binge drinking (RR 1.5, 95% CI: 1.3-1.7) and frequent drinking (RR 1.4, 95% CI: 1.3-1.5) increasing during the pandemic, while risk of having 5+ drinks on a typical drinking day was reduced (RR 0.4, 95% CI: 0.3-0.5). There were also statistical interactions between time period and age group, as well as time period and gender for all alcohol outcomes and with education level for binge drinking (Tables S5-7).

### Cigarette smoking and e-cigarette use

Current cigarette smoking decreased during lockdown (Figure 1 and Table S2). The decrease in smoking was more apparent in younger age groups and among men (Figure 3) and seems driven by a decline in lighter smokers (Table S2). Longitudinal models demonstrated that risk of smoking reduced during the pandemic (RR 0.9, 95% CI: 0.8-1.0) (Table 2), but there were no statistically significant (p<0.05) interactions with age group, gender, race/ethnicity or education level (Table S8). In the longitudinal analyses, risk of e-cigarette use was also lower during the pandemic (RR 0.7, 95% CI: 0.5-0.9) (Table 2), but no statistically significant interactions were found with the subgroups examined (Table S9).

## Discussion

Psychological distress substantially increased in the UK following the COVID-19 pandemic. Groups most adversely affected included women and younger people. The increase in psychological distress, measured after the first month of lockdown, appeared to be driven by a reduction in enjoyment of normal day-to-day activities, as well as increased difficulties with concentration and sleep, and feelings of unhappiness. Overall, loneliness remained relatively stable. Cigarette smoking declined, and this reduction appears to reflect cessation among lighter smokers. The frequency of drinking four or more times a week and binge drinking increased, particularly among those over 25 and white people.

Our study has several strengths. We used a large nationally representative longitudinal dataset. We also checked the variation in our outcomes before the pandemic and found that secular trends tended to be small compared to changes observed during the pandemic. Some limitations should be noted. First, survey non-participation may have introduced bias in our estimates, especially as the response rate in the COVID survey was lower than usual. However, weights were used to reduce concerns about non-response and attrition. Second, there were changes in the modality by which the COVID survey was administered (moving from mixed mode (face-to-face, web and phone) to online surveys)), which may have led to modest reporting changes. However, empirical investigation suggested this is unlikely to have biased responses.^22^ Relatedly, there were minor changes to the questionnaire items about alcohol consumption, so that questions related to the pandemic period rather than the entire previous year. This meant that a modified version of the AUDIT-C scale was used, which is not strictly comparable with previous years and so these initial results should be interpreted with caution. Alcohol consumption is also known to be under-reported in surveys.^23 24^ The pandemic context may have also influenced participant reporting more broadly. For example, the increase in being less able to enjoy usual activities may not reflect anhedonia, but rather the reality of experiencing lockdown and could be considered a normal response. Relatedly, what people perceive as a ‘typical’ drinking day is likely to have changed, especially among younger people, which could explain the conflicting results for this measure of alcohol consumption.

While a body of literature is developing to articulate the expected indirect impacts of the pandemic^3^ ^7 25^, empirical research on how mental health and health-related behaviours have changed remains limited and largely based on non-representative samples.^26^ A repeated cross-sectional analysis comparing results of two different representative surveys conducted before and after the pandemic in the USA found a marked increase in psychological distress amongst adults, from 3.9% to 13.6%.^27^ Furthermore, younger people experienced the greatest relative increase in poor mental health, echoing our findings. While longitudinal evidence on changes in consumption of tobacco and alcohol are limited, some cross-sectional surveys have been conducted which asked about self-perceived changes in behaviour. A representative survey conducted on behalf of the charity Alcohol Change UK found that 21% of adults who normally drink alcohol self-reported increased consumption, but 35% reduced how often they drink or have stopped drinking altogether.^28^ Similarly, an online non-representative survey with data collection following the pandemic, also found that self-reported tobacco and e-cigarette use reduced by about one-quarter.^29^

Our study has important implications for public health policy. The substantial increase in psychological distress in the UK highlights the potential tension between implementing lockdown measures to control the pandemic and the risk of health harms that such action could have. By comparison, in England poor mental health after the Great Recession (assessed using the same GHQ outcome used in this study) increased from 13.7% to 16.4%^30^ an effect size approximately one-quarter of that observed in this study. Finding that women have been disproportionately affected illustrates broader unequal power relations within society, with women more likely to experience the additional burden of childcare and more likely to work in sectors worst affected by the pandemic.^31^ It is worth noting that this more recent decline in mental health among women occurs after a period of austerity, during which women’s mental health had already been showing adverse trends.^32-34^ The reduction in smoking, despite the adverse societal circumstances, may illustrate the importance of the availability of these products in influencing behaviour. There is an increasing evidence base which suggests that availability of unhealthy commodities drives consumption and contributes to health inequalities.^35^ The trends in alcohol consumption merit further exploration. The frequency of alcohol consumption and binge drinking appear to have increased, but the proportion of people drinking 5+ drinks on a typical day when drinking decreased. This may reflect change in what a typical drinking day is (e.g. going to the pub with friends compared to drinking at home) and the change in the frequency of alcohol consumption.

Further research is needed to understand mechanisms by which these impacts may be arising and whether the large increase in psychological distress remains following changes to the lockdown. As we did not investigate differences by country of the UK, future research should investigate whether there are differential trends over the longer-term course of the pandemic by country as the mitigation policies began to diverge. We also found psychological distress increased among the most educated groups, which may reflect that this group was more likely to move to remote working during the pandemic, and for some, this was combined with the home-schooling of children. Monitoring this group to see if they are better able to recover from the initial shock of the lockdown will be important to understand the implications for mental health inequalities. While the UK Government introduced aggressive fiscal policies to minimise adverse economic risks, it is likely that at least some of these impacts reflect the start of a potentially long-lasting economic crisis.^36^ Understanding to what extent health is also being impacted by income and unemployment shocks will help inform decisions about ongoing support over the coming months and years.^37^ However, improved psychological support, including access to mental health services, may also be necessary. Our research provides an early picture of the broader consequences of the pandemic – clearly, longer term monitoring will be necessary. Poor mental health is an important predictor of future mortality and several physical health conditions.^38 39^ Given this, further monitoring of the determinants of health, as well as health outcomes, are required.

## Data Availability

Understanding Society deidentified survey participant data are available through the UK Data Service (http://doi.org/10.5255/UKDA-SN-6614-13; http://doi.org/10.5255/UKDA-SN-8644-3). Researchers who would like to use Understanding Society need to register with the UK Data Service (https://ukdataservice.ac.uk/) before being allowed to download datasets.

## Acknowledgements

We would like to thank the participants of the Understanding Society study. The Understanding Society COVID-19 study is funded by the Economic and Social Research Council (ES/K005146/1) and the Health Foundation (2076161). Fieldwork for the survey is carried out by Ipsos MORI and Kantar. Understanding Society is an initiative funded by the Economic and Social Research Council and various Government Departments, with scientific leadership by the Institute for Social and Economic Research, University of Essex.

## Authors’ contributions

SVK and CLN conceived the idea for the study. CLN, MJG and SVK conducted the analysis. SVK, CLN and MJG drafted the manuscript. CLN, MJG, MB, DDC, PC, ED, AHL, AP, RMT, EW and SVK contributed to the study design, interpretation of the findings and critically revised the manuscript. All authors approved the final version of the paper. CLN, MJG and SVK had full access to the study datasets and act as guarantors.

## Licence for publication

The Corresponding Author has the right to grant on behalf of all authors and does grant on behalf of all authors, an exclusive licence (or non exclusive for government employees) on a worldwide basis to the BMJ Publishing Group Ltd to permit this article (if accepted) to be published in JECH and any other BMJPGL products and sublicences such use and exploit all subsidiary rights, as set out in our licence (http://group.bmj.com/products/journals/instructions-for-authors/licence-forms).

